# A variational sparse Gaussian-process method for detecting spatially variable genes and cellular interactions from spatial transcriptomics

**DOI:** 10.64898/2025.12.10.25341956

**Authors:** Zhicong Wang, Liqing Xie, Yiran Wang, Yongtian Wang, Tao Wang, Xuequn Shang, Jing Li, Jialu Hu

## Abstract

Advanced spatially resolved transcriptomic (SRT) technologies preserve the spatial context of gene expression within tissues, enabling the study of context-dependent transcriptional regulation. Here, we propose VISGP, a variational sparse gaussian-process method for spatial variable genes (SVGs) and cellular interactions analysis from such data. VISGP utilizes variational inference and a sparse Gaussian process approximation, which efficiently models the posterior distribution with a set of inducing variables, thereby minimizing computational and memory complexity. When applied to simulated data and four real data sets, VISGP successfully identified the most SVGs than existing methods, and detected 85 spatially constrained ligand–receptor pairs that are missed by other methods. Together, VISGP provides a powerful strategy for decoding spatial gene regulation and cellular interactions, offering valuable biological insights into cellular heterogeneity and cancer pathology.

**Teaser:** A statistical framework that uncovers spatially variable genes and cell-cell communication, revealing hidden biological architecture in tissues.

## Introduction

Spatially resolved transcriptomics (SRT) enables direct measurement of gene expression within intact tissue architecture, transforming analyses of cellular heterogeneity. By preserving spatial context, these technologies have yielded fundamental insights into tissue function (*1, 2*), normal development (*3*), and disease pathology (*4*). Over the past decade, diverse SRT technologies have emerged, differing in resolution, sensitivity, and transcriptome coverage (*5–7*). Imaging-based techniques, such as in situ sequencing (ISS) and multiplexed in situ hybridization methods (e.g., MERFISH and seqFISH), provide single-molecule resolution but are limited to predefined target panels. In contrast, sequencing-based methods encode spatial information onto RNA molecules and recover transcriptome-wide expression through next-generation sequencing (*8*). The growing scale of these datasets requires computational frameworks to resolve spatial heterogeneity and uncover tissue architecture. A key analytical goal in SRT is to identify spatially variable genes (SVGs), which differ from highly variable genes in scRNA-seq. SVGs show region-specific expression patterns that mirror tissue organization and cellular heterogeneity, providing a quantitative link between transcriptional variation and anatomical structure.

Several statistical modeling methods have been proposed to identify significant SVGs based on various model-based inference. The most pioneering methods include trendsceek (*9*), SpatialDE (*10*), SPARK (*11*), and SPARK-X (*12*). A more recent method, nnSVG (*13*), employs a nearest-neighbor Gaussian process (NNGP) framework to estimate gene-specific spatial covariance parameters. However, these model–based approaches often rely on predefined kernel functions with empirically assigned hyperparameters, resulting in a shared covariance structure across genes that may not adequately capture diverse spatial expression patterns.

Apart from statistical modeling, several deep learning methods have been developed for SRT data, including SOMDE (*14*), SpaGCN (*15*), SPADE (*16*) and STAMarker (*17*). These techniques usually construct spatial dependency structures first, treating graph generation, constraint design, or normalization as a separate module. Any inaccuracies introduced during this initial step tend to be inherited by downstream analyses, potentially leading to less optimal outcomes. Collectively, these limitations constrain the robustness and generalizability of current approaches in detecting spatially variable gene expression.

Here, we present VISGP (Variational Inference–assisted Sparse Gaussian Process), a new statistical modeling framework for identifying SVGs. VISGP models each gene’s expression as a Gaussian process integrating sparse approximation with variational inference (*19–21*) to achieve both flexibility and computational efficiency. The sparse formulation introduces a set of low-dimensional inducing variables that compress spatial features and reduce computational complexity, while variational inference jointly optimizes the kernel hyperparameters with model parameters, enabling gene-specific adaptation of spatial covariance structures. Furthermore, it employs a score-based statistical test under a null-hypothesis framework (*22*) to rigorously identify SVGs. We benchmarked VISGP against three commonly used methods on one simulated and three real SRT datasets. It accurately and efficiently detected spatially variable genes, demonstrating strong robustness and scalability. Compared with existing methods, it exhibited superior sensitivity in capturing subtle spatial patterns, particularly in datasets with weak spatial signals. Beyond identifying spatially variable genes, it can also be readily extended to infer spatially dependent ligand–receptor interactions, enabling characterization of spatially organized cell–cell communication. These results highlight the potential of VISGP as a powerful and generalizable tool for the analysis of SRT data.

## Results

### Methods overview

As illustrated in Fig. 1a-d, VISGP integrates gene expression profiles and spatial coordinates to detect spatially variable genes from SRT data. For each gene, we model expression as a Gaussian process with a squared exponential (RBF) kernel parameterized by length scale *ℓ* and signal variance *τ*, and assume Gaussian noise with variance 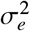. All model and variational parameters are jointly optimized by maximizing the *evidence lower bound (ELBO)* under a sparse variational approximation with inducing variables (*19–21*). The inducing locations **Z** and variational parameters (***μ*, Σ**) are learned together with the model parameters, enabling scalable inference with time complexity O(*nm*^2^) and memory O(*nm*) for *n* spots and *m* inducing points. We then perform a *score-based test* (*22*) under the null hypothesis *H*_0_ : *τ* = 0 to call spatially variable genes, where the test statistic *Q* = **y**^*T*^ **K**_*nn*_**y** follows a mixture of *χ*^2^ distributions (Fig. 1c). Genes passing the significance threshold are visualized together with their spatial expression patterns (Fig. 1d). In practice, we apply a Gaussian likelihood after library-size normalization and log-transformation of counts, which yields approximately homoscedastic residuals suited to GP modeling. An implementation is available in our Python package scbean (version ≥ 0.5.0; see Code availability).

**Figure 1:**
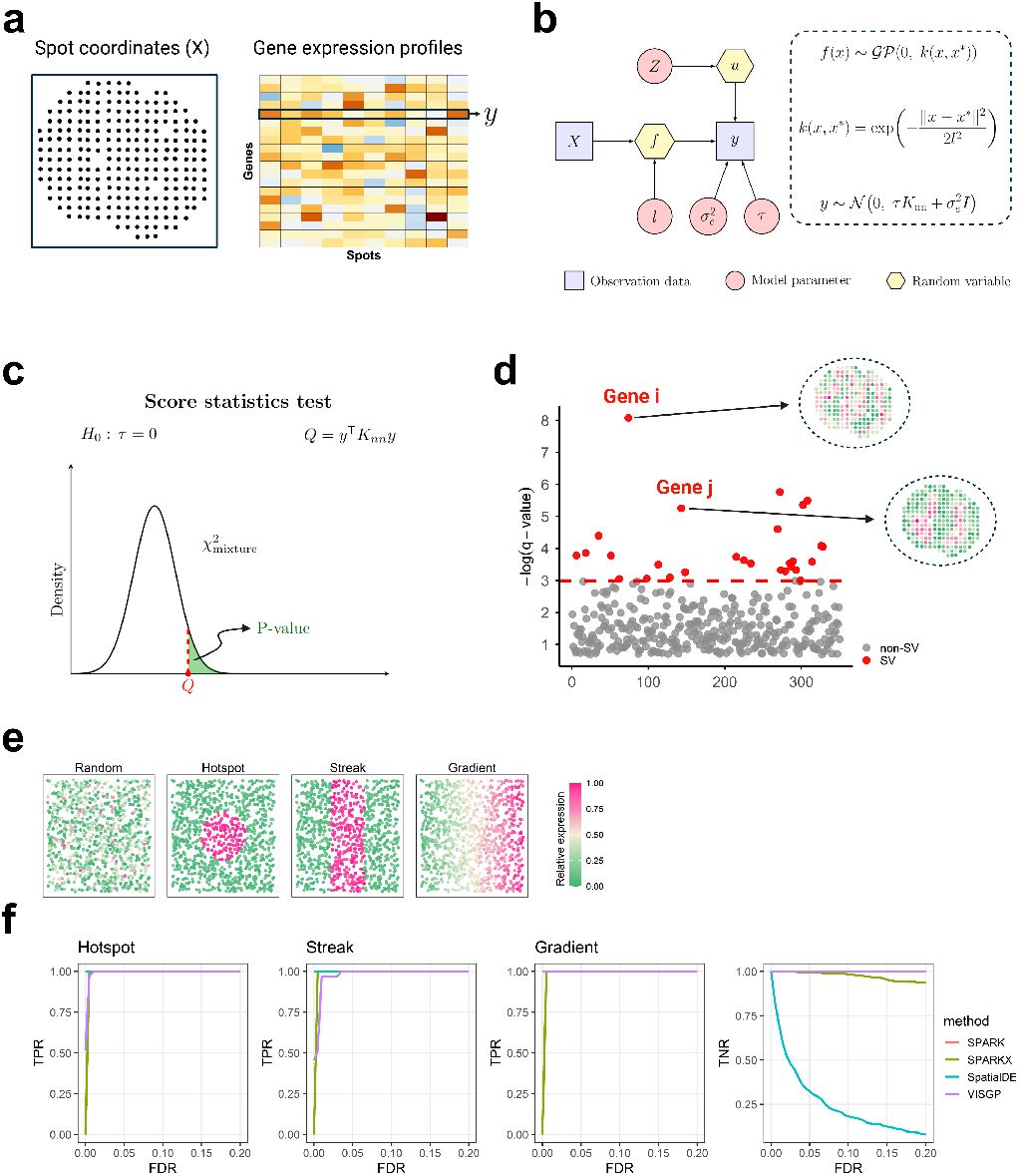
Method overview of VISGP and simulation results. (a) Model inputs. (b) Variational Inference assisted Sparse Gaussian Process. **K**_*nn*_ is a covariance matrix whose elements are given by the kernel function *k* (*x, x*^∗^) for each pair of spot coordinates. (c) Score statistical significance test. SV genes are screened by q-value (adjusted p-value). The red dotted line represents the q-value cutoff of 0.05. (e) Representative genes in the set of simulation show a random pattern and other three spatial expression patterns (hotspot, streak, gradient). (f) Power plots illustrate the proportion of true positives rate identified and true negatives rate identified by different methods across a range of FDRs in the set of simulation.

### Performance assessment on simulated data

To benchmark VISGP against current state-of-the-art methods, including SpatialDE, SPARK, and SPARK-X, we applied each algorithm to one set of simulation. This simulation was designed to evaluate how accurately the methods distinguish SVGs under controlled spatial patterns. Following Trendsceek (*9*), we generated spatial pseudo-count data for the simulation. Spatial coordinates for 1,000 spots were sampled from a Poisson point process, and gene expression counts were drawn from a negative binomial distribution. Non-spatially variable genes (non-SVGs) were randomly distributed without spatial architecture, whereas SVGs exhibited one of three spatial patterns— hotspot, streak, or gradient (Fig. 1e). Each simulated dataset contained 400 non-SVGs and 100 SVGs representing these distinct spatial configurations. For a fair comparison, we evaluated all methods using sensitivity (true positive rate, TPR), specificity (true negative rate, TNR), and false discovery rate (FDR) as performance metrics.

In terms of sensitivity, all four methods accurately identified SVGs across the different spatial patterns (hotspot, streak, or gradient) at conservative thresholds (FDR *<*10%). Regarding specificity, SpatialDE showed a marked decline as the FDR threshold increased, this reduction likely reflects limitations of its spatial kernel in modeling certain expression patterns. While SPARK-X exhibited a mild decrease, however, at an FDR of 0.05, the reduction in specificity for SPARK-X was minimal (Fig. 1f). Overall, VISGP and SPARK effectively distinguished SVGs from non-SVGs in the set of simulation, whereas SpatialDE tends to misclassify some non-SVGs as SVGs. These findings provide a solid foundation for applying VISGP to real SRT datasets to discover biolgoical structure with different spatial patterns.

### VISGP enables robust SVGs analysis in the mouse olfactory bulb data

We next applied VISGP to a real SRT dataset of the mouse olfactory bulb (MOB) (*23*), comprising 15,284 genes measured across 237 spots (see *Dataset and Preprocessing* for details). As benchmarks, SpatialDE, SPARK, and SPARK-X were applied with their recommended configurations. To obtain a null expectation, we kept the real MOB expression matrix unchanged and randomly permuted the spatial coordinates 10 times. Each permuted coordinate file was analyzed with these four methods. We found that VISGP, SPARK, and SPARK-X yielded well-calibrated p-values under the permuted null, whereas SpatialDE showed pronounced p-value deflation (Supplementary Fig. 1a). We next evaluated performance on the real (unpermuted) spatial data, as shown in Fig. 2a–b, VISGP recovered 1,607 SVGs at an FDR of 0.05, outperforming SPARK-X and SPARK by nearly twofold and SpatialDE by fivefold. Among these, VISGP, SpatialDE, SPARK-X, and SPARK uniquely reported 679, 5, 137, and 402 model-specific SVGs, respectively. We subsequently grouped the 1,607 SVGs identified by VISGP into three principal categories (Fig. 2c,d), each representing a distinct spatial expression pattern: Pattern I, Pattern II, and Pattern III. In the mouse olfactory bulb, Pattern I marked genes enriched in the glomerular layer, Pattern II in the mitral cell layer, and Pattern III in the granule cell layer, mirroring the region’s hierarchical neuronal organization. These three patterns were further validated by representative marker genes, including Penk (*p <* 1 × 10^−16^), Doc2g (*p* ≤ 5.3 × 10^−3^), and Kctd12 (*p <* 1 × 10^−16^) (Fig. 2e). To assess cross-method consistency, model-specific SVGs uniquely identified by VISGP, SPARK, and SPARK-X were also clustered into three categories. The spatial patterns defined by VISGP- and SPARK-specific genes remained consistent with the three anatomical layers of the MOB (Supplementary Fig. 2a,b), whereas those from SPARK-X were less well-defined (Supplementary Fig. 2c). Corresponding heatmaps of VISGP-specific SVGs revealed well-defined spatial expression patterns (Supplementary Fig. 2d). Furthermore, we evaluated the spatial coherence of SVGs uniquely identified by each method using Moran’s I, a standard statistic that quantifies spatial autocorrelation on a scale from –1 (complete dispersion) to 1 (perfect clustering). VISGP-specific SVGs exhibited higher Moran’s I values compared to those identified by SPARK and SPARK-X, indicating stronger spatial organization and biological relevance (Supplementary Fig. 2e). We further examined the biological significance of the 1,607 SVGs uniquely identified by VISGP using gene set enrichment analysis (GSEA). A total of 756 Gene Ontology (GO) terms were significantly enriched (Benjamini– Hochberg adjusted *p* ≤ 0.05). As shown in Fig. 2f and Supplementary Fig. 3, the most prominent enriched categories were related to neuronal and synaptic processes, including “neuron-to-neuron synapse” (GO:0098984; adjusted *p* = 3.18 × 10^−23^) and “asymmetric synapse” (GO:0032279; adjusted *p* = 5.92 × 10^−22^). Additional terms such as “modulation of chemical synaptic transmission” (GO:0050804; adjusted *p* = 6.75 × 10^−20^) and “regulation of synapse organization” (GO:0050803; adjusted *p* = 3.90 × 10^−14^) further highlight the enrichment of neuronal communication pathways. These findings indicate that VISGP effectively captures spatial gene expression patterns associated with neuronal circuitry, consistent with the known layered organization of the olfactory bulb.

**Figure 2:**
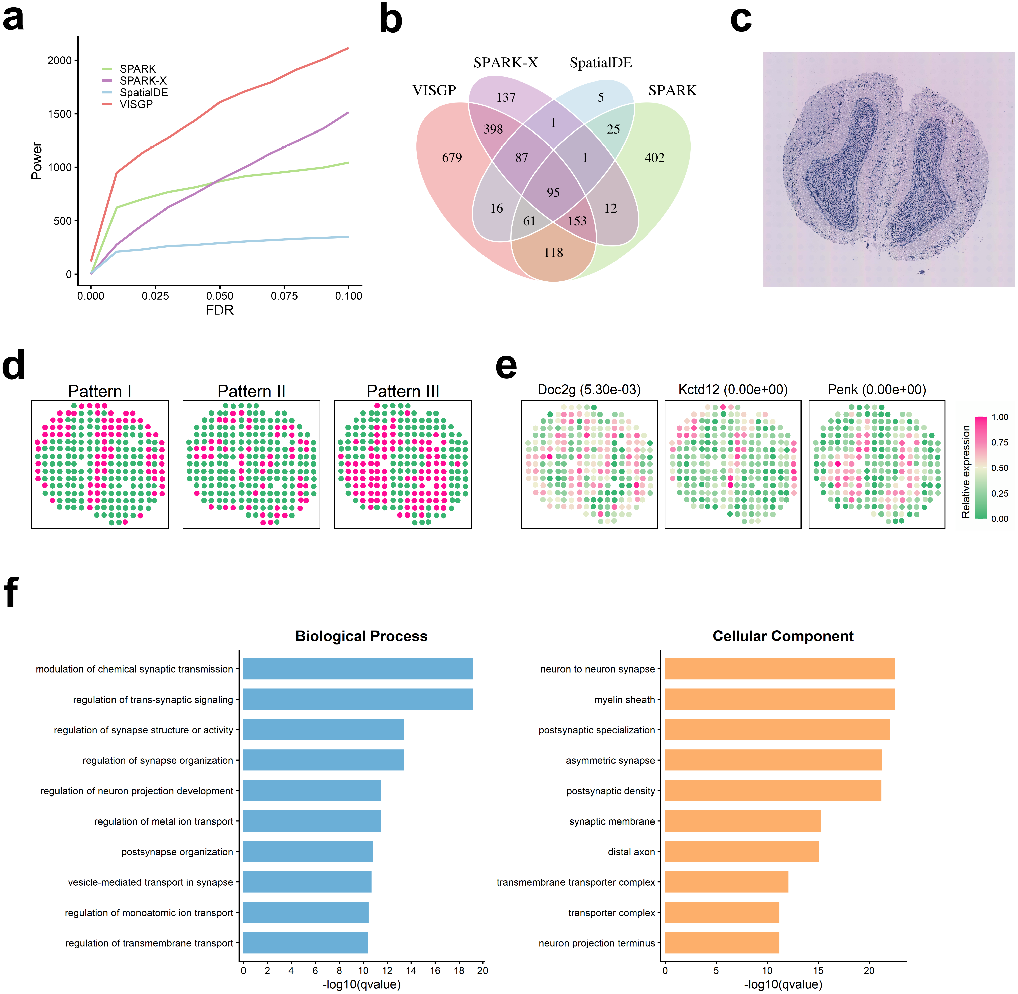
Analyzing the mouse olfactory bulb dataset. (a) Power plot shows the number of SV genes (y-axis) detected by different methods in a series of FDRs in this data. (b) Venn diagram shows the overlap between SV genes identified by the four methods based on an FDR cutoff of (c) Image of hematoxylin and eosin-stained mouse olfactory bulb from (*23*). (d) According to the SV genes identified by VISGP, three different spatial expression patterns are summarized. (e) Spatial expression pattern for some marker genes for different layers in mouse olfactory bulb along with their FDR (adjusted p-value) from VISGP (in parentheses). Color represents relative gene-expression levels (pink, high; green, low). (f) Bar plot for GO enrichment analysis on 1607 SV genes obtained by VISGP based on an FDR cutoff of 0.05. Gene sets are colored by two categories: GO biological process (green) and GO cellular component (orange).

### VISGP reveals biologically meaningful structure in the human breast cancer microenvironment

To evaluate VISGP’s performance in tumor tissues, we applied it to a SRT dataset of human breast cancer biopsies (*23*), comprising 14,789 genes measured across 251 spots. The same dataset was also analyzed using SpatialDE, SPARK, and SPARK-X for comparison. Permutation analysis revealed that VISGP, SPARK, and SPARK-X maintained proper p-value calibration under the null, whereas SpatialDE exhibited systematic p-value inflation, deviating upward from the theoretical null (Supplementary Fig. 1b). We next proceeded to evaluate the methods on the real spatial data, VISGP exhibited higher sensitivity across FDR thresholds ranging from 1%–10% (Fig. 3a). At an FDR of 5%, VISGP identified 3,302 SVGs, compared with 2,032 from SPARK-X, 469 from SPARK, and 201 from SpatialDE. Among these, 1,552 SVGs were uniquely detected by VISGP, 306 by SPARK-X, 28 by SPARK, and 39 by SpatialDE (Fig. 3b). We then clustered the 3,302 SVGs identified by VISGP into two dominant spatial expression patterns (Fig. 3d). Pattern I was associated with the cancer tissue region visually depicted as the dark area in Fig. 3c, whereas Pattern II mapped to adjacent non-tumorous tissue. This spatial contrast highlights VISGP’s ability to distinguish between biologically distinct regions within tumor sections. To assess biological validity, we benchmarked VISGP its competitors using a curated list of 18 previously reported tumor-associated genes (*23–25*). As shown in Fig. 3e and Supplementary Fig. 4, VISGP identified all 18 genes with well-defined spatial expression patterns, whereas SpatialDE, SPARK, and SPARK-X detected 6, 10, and 12 of them, respectively. To further evaluate the biological relevance of VISGP-specific discoveries, we analyzed the 1,552 SVGs uniquely identified by VISGP. Clustering revealed two distinct spatial expression patterns, both exhibiting clear spatial organization. In contrast, SVGs uniquely identified by SPARK and SPARK-X showed weaker and less defined spatial patterns (Supplementary Fig. 5). We next performed Gene Ontology (GO) enrichment analysis on both clusters of SVGs to investigate their functional significance. At an FDR threshold of 0.05, 370 GO terms were significantly enriched (Fig. 3f). Among them, “cytoplasmic translation” (GO:0002181; *p* = 3.13 × 10^−21^) and “cytosolic ribosome” (GO:0022626; *p* = 8.26 × 10^−23^) were the most enriched, consistent with enhanced protein synthesis activity in tumor cells—a hallmark supporting rapid growth and metabolic reprogramming. We further examined pathway enrichment by spatial pattern (Supplementary Fig. 6). Pattern I genes were associated with “extracellular matrix organization” (GO:0030198; *p* = 7.08 ×10^−17^), “focal adhesion” (GO:0005925; *p* = 1.72 ×10^−19^), and “blood vessel morphogenesis” (GO:0048514; *p* = 4.99 ×10^−10^), reflecting active ECM remodeling, angiogenesis, and cell–matrix interactions within the tumor microenvironment. In contrast, Pattern II genes were enriched in “oxidative phosphorylation” (GO:0006119; *p* = 1.14 × 10^−12^) and “ATP synthesis coupled electron transport” (GO:0042773; *p* = 4.85 × 10^−12^), suggesting preserved mitochondrial metabolism in adjacent non-tumorous regions. In conclusion, these analyses demonstrate that VISGP couples high-sensitivity detection of spatially variable genes with clear delineation of biologically meaningful tumor microenvironment structure.

**Figure 3:**
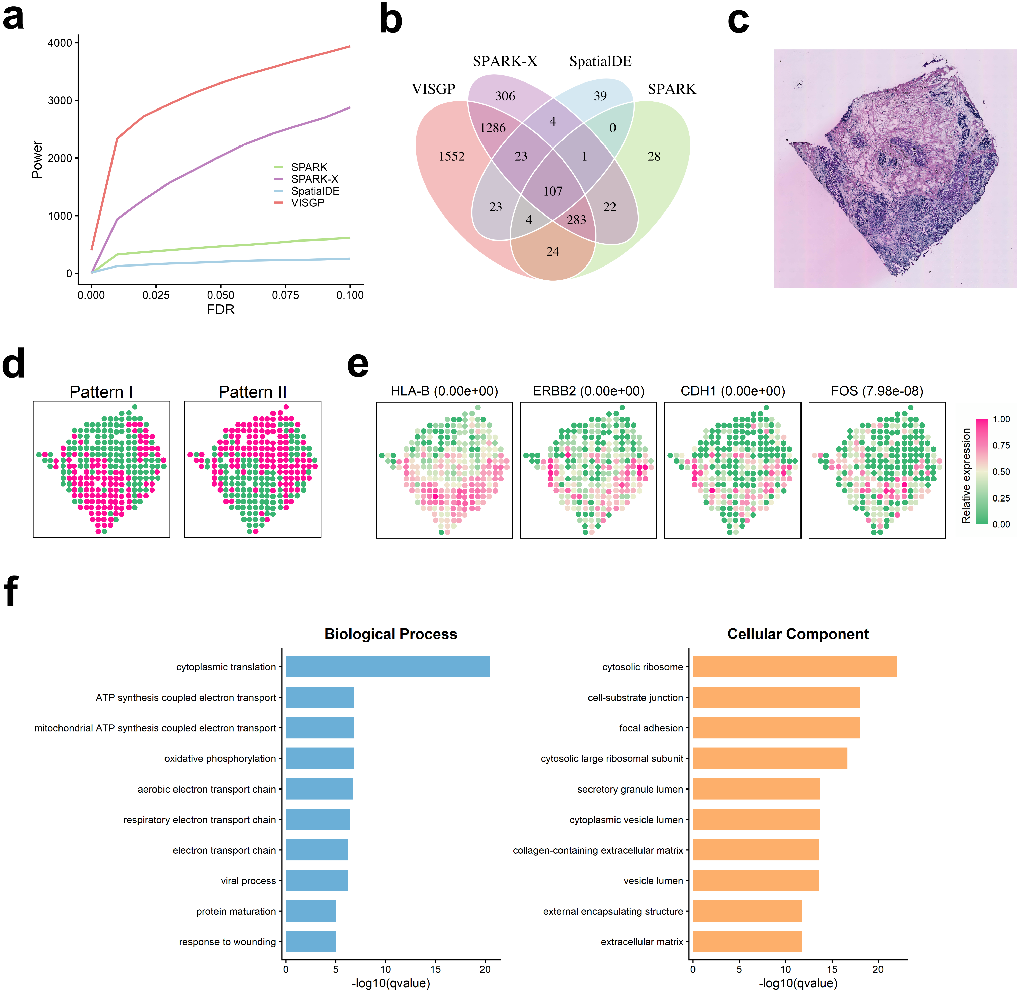
Analyzing the human breast cancer dataset. (a) Power plot shows the number of SV genes detected by different methods in a series of FDRs in this data. (b) Venn diagram shows the overlap between SV genes identified by the four methods based on an FDR cutoff of 0.05. (c) Image of hematoxylin and eosin-stained human breast cancer from (*23*). (d) According to the SV genes identified by VISGP, two different spatial expression patterns are summarized. (e) Spatial expression pattern for four known tumor genes. The FDRs for the four genes from VISGP are shown inside parentheses. Color represents relative gene-expression levels (pink, high; green, low). (f) Bar plot for GO enrichment analysis on 3302 SV genes obtained by VISGP based on an FDR cutoff of 0.05. Gene sets are colored by two categories: GO biological process (green) and GO cellular component (orange).

### VISGP detects unique spatial patterns in human squamous cell carcinoma

In this third application, we applied VISGP to a human cutaneous squamous cell carcinoma dataset from a left forearm specimen (*26*). In total, 16,383 genes were profiled across 521 spots. Consistent with breast cancer data, VISGP, SPARK, and SPARK-X showed proper null calibration under permutated null, whereas SpatialDE did not (Supplementary Fig. 1c). On the real spatial data, VISGP identified 3,213 SVGs at an FDR of 0.05, followed by SPARK-X (3,044), whereas SpatialDE detected substantially fewer (216), indicating lower sensitivity relative to the other methods (Fig. 4a-b). We assessed spatial autocorrelation of the SVGs identified by the three methods using Moran’s I. VISGP yielded consistently higher Moran’s I values than SPARK-X and SpatialDE (Supplementary Fig. 7a), indicating stronger spatial architecture in the detected genes. When restricted to SVGs uniquely identified by each method (Supplementary Fig. 7b), VISGP still produced higher Moran’s I scores, confirming that its discoveries greater spatial coherence. Several key genes highlighted in the original study were also recovered by VISGP (Fig. 4d; Supplementary Fig. 8a–c), including tumor-associated, immunosuppression-related, and T cell–marker genes. Notably, VISGP uniquely identified additional SVGs that revealed biologically relevant patterns not captured by the other methods (Supplementary Fig. 8d). For example, *TNC, ITGA5*, and *LAMB2* delineate an extracellular matrix (ECM)–integrin axis underlying fibroblast-driven matrix remodeling and invasion. Recent studies have synthesized *TNC*’s matricellular roles in mechanical and immune niches and discussed emerging TNC-targeting strategies, while *ITGA5* and *LAMB2* have been associated with pro-invasive and prognostic programs across cancers (*27–29*). In addition, *CSF1R* and *ENG* were uniquely detected by VISGP as spatially variable genes, revealing distinct myeloid-enriched and angiogenic niches within squamous carcinoma tissues—findings consistent with recent evidence showing that myeloid remodeling can drive resistance to CSF1R inhibition (*30*) and that ENG acts as a key endothelial regulator of VEGF-driven angiogenic sprouting (*31*). We next performed Gene Ontology (GO) enrichment analysis on the SVGs identified by VISGP. A total of 335 GO terms were significantly enriched (Fig. 4e). Terms such as “cytoplasmic translation” (GO:0002181; *p* = 1.37 × 10^−28^) and “cytosolic ribosome” (GO:0022626; *p* = 4.61 × 10^−25^) were highly enriched, indicating enhanced ribosomal biogenesis and protein synthesis consistent with the high proliferative activity of malignant keratinocytes. Processes including “epidermis development” (GO:0008544; *p* = 1.06 × 10^−8^), “skin development” (GO:0043588; *p* = 3.03 × 10^−8^), and “keratinocyte differentiation” (GO:0030216; *p* = 1.43 × 10^−6^) reflected the preservation of epithelial lineage programs within SCC tissue structure. In addition, enrichment of “cell–substrate junction” (GO:0030055; *p* = 1.23 × 10^−18^) and “focal adhesion” (GO:0005925; *p* = 3.11 × 10^−18^) terms indicated active cell–matrix interactions, a hallmark of invasive SCC fronts. Together, these results demonstrate that VISGP captures spatially organized transcriptional programs integrating protein synthesis, epithelial differentiation, and matrix adhesion in squamous carcinoma.

**Figure 4:**
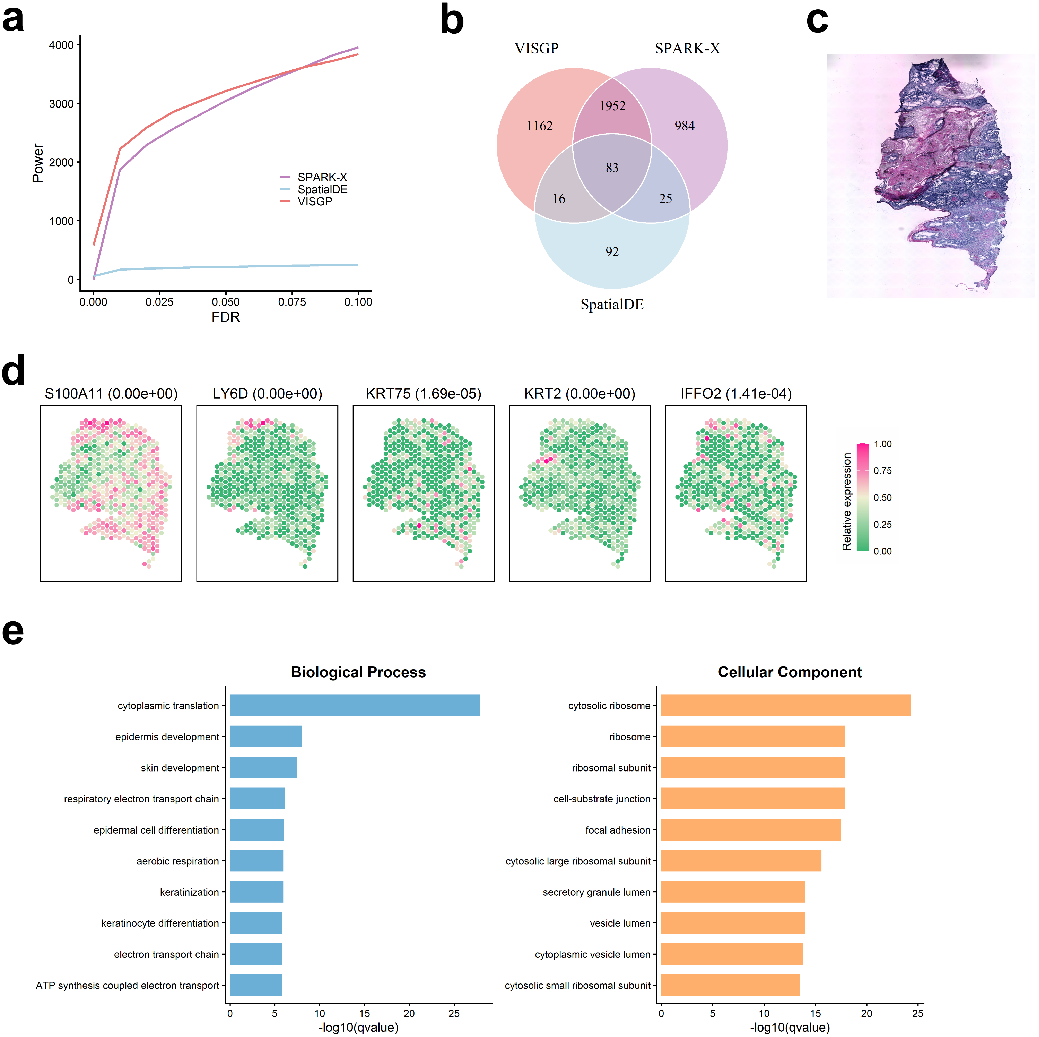
Analyzing the human squamous cell carcinoma dataset. (a) Power plot shows the number of SV genes detected by different methods in a series of FDRs in this data. (b) Venn diagram shows the overlap between SV genes identified by the three methods based on an FDR cutoff of 0.05. (c) Image of hematoxylin and eosin-stained human squamous cell carcinoma. (d) Spatial expression pattern for some representative genes. The first three genes (S100A11, LY6D, KRT75) are associated with tumors, and the last two (KRT2, IFFO2) are associated with non-tumor adjacent stroma. The FDRs for the five genes from VISGP are shown inside parentheses. Color represents relative gene-expression levels (pink, high; green, low). (e) Bar plot for GO enrichment analysis on 10,828 SV genes obtained by VISGP on the basis of an FDR cutoff of 0.05. Gene sets are colored by two categories: GO biological process (green) and GO cellular component (orange).

### VISGP reveals spatially constrained ligand-receptor interactions in human pancreatic ductal adenocarcinoma

To evaluate the performance of VISGP, we applied it to a SRT dataset of human pancreatic ductal adenocarcinoma (PDAC) (*32*). The dataset comprised multiple histologically annotated regions— including cancerous, pancreatic, ductal, and stromal areas—based on H&E staining, and contained expression profiles of 25,754 genes across 428 spatially resolved spots. We modeled the spatial expression of each gene in a curated ligand–receptor database (*33*) using VISGP and quantified the spatial similarity between ligand–receptor pairs using an *asymmetric* Kullback–Leibler (KL) divergence measure. Statistically significant ligand–receptor interactions were determined by permutation testing of KL values (Fig. 5a). We identified 95 ligand–receptor pairs showing statistically significant spatially coordinated expression (*p <* 0.05). Spatial heatmaps revealed co-localized or adjacent expression domains for these pairs, particularly for *TIMP1*–*CD63* (Fig. 5b), with additional examples provided in Supplementary Fig. 9. *TIMP1*, a tissue inhibitor of metalloproteinases, has been linked to poor prognosis in multiple cancers, including PDAC (*34*). Its interaction with *CD63* promotes the formation of neutrophil extracellular traps (NETs) within the tumor microenvironment, contributing to immune evasion and metastatic potential (*35*). Beyond the *TIMP1*–*CD63* axis, VISGP revealed additional PDAC-relevant ligand–receptor pairs. *LAMC2*–*ITGB4* marked laminin–integrin adhesion tracks at invasive tumor fronts, where laminin-332 engagement with integrin *β*4 reinforces epithelial–stromal attachment and facilitates epithelial–mesenchymal transition, promoting collective invasion (*36, 37*). *SEMA3F*–*PLXNA3* represented a class-3 semaphorin signaling axis mediating tumor–nerve interactions; binding of *SEMA3F* to *PLXNA3* activates axon-guidance–like pathways that support neural remodeling and perineural invasion within the desmoplastic stroma (*38*). To assess the added value of spatial modeling, we compared the VISGP-inferred ligand–receptor interactions with those derived from a matched single-cell RNA-seq dataset of the same patient (PDAC-A) (*32*) analyzed using the CellPhoneDB pipeline (*39*). The approach identified 156 ligand–receptor pairs across diverse cell populations such as acinar cells, Ductal– MHC–Class II cells, Ductal–CRISP3–high centroacinar-like cells, and mDCs. However, none of the four representative ligand–receptor pairs detected by VISGP (Fig. 5b) were recovered by the scRNA-seq analysis. Among the 95 ligand–receptor pairs identified by VISGP, only 10 overlapped with those from CellPhoneDB, revealing that VISGP captured 85 spatially constrained and tissue-specific communication events not detectable by conventional scRNA-seq–based inference. These findings demonstrate that incorporating spatial information substantially enhances the discovery of context-dependent signaling interactions within the tumor microenvironment. Pathway enrichment analysis showed that the VISGP-identified ligand–receptor pairs were significantly associated with key cancer-related signaling pathways, including PI3K–Akt, ECM–receptor interaction, cytokine– cytokine receptor interaction, and focal adhesion (Fig. 5d). Among these, the ECM–receptor interaction and focal adhesion pathways exhibited strong spatial dependence, as they rely on direct cell–matrix contact and mechanical coupling at the tumor–stroma interface. In contrast, cytokine– cytokine receptor signaling displayed moderate spatial dependence through short-range paracrine gradients within immune and fibroblast compartments, supporting tumor-promoting crosstalk in the local microenvironment (*40*). The PI3K–Akt pathway, although primarily intracellular, acted as a downstream effector of these spatially anchored signaling axes and is known to be hyperactivated in pancreatic cancer, contributing to apoptosis resistance and therapeutic evasion (*41*).

**Figure 5:**
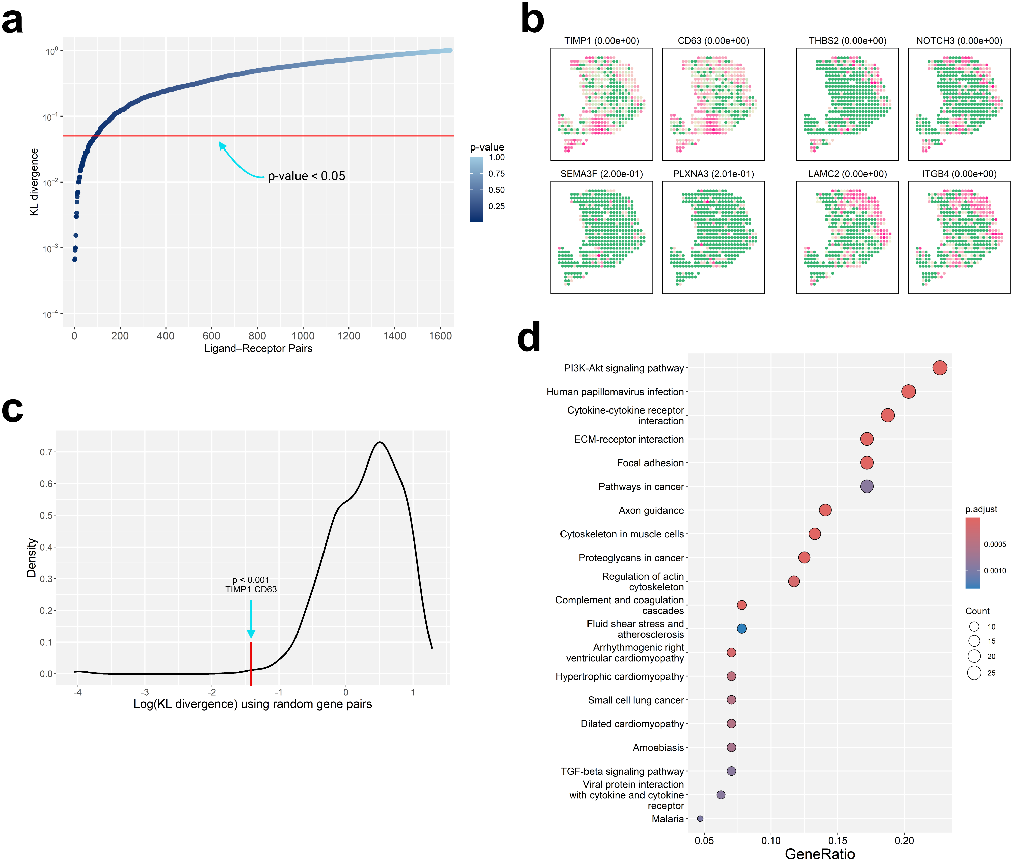
Analysis results of ligand-receptor pairs predicted by VISGP. (a) Scatter plot shows the ligand-receptor pairs and their corresponding KL divergence. The color of the points represent the p-value. (b) Heatmaps of four ligand-receptor pairs that have similar expression distribution in space. (c) The distribution of KL divergence of randomly sampled gene pairs. We performed the statistical test on this background distribution. (d) KEGG enrichment shows that ligand-receptor pairs predicted by VISGP are closely related to human PDAC.

## Discussion

Spatial transcriptomics enables quantitative dissection of gene expression within intact tissue structure (*1, 2, 5*); however, extracting biologically meaningful spatial organization remains a key analytical challenge (*10–12*). In this study, we developed VISGP, a variational inference–assisted sparse Gaussian process framework that achieves scalable, gene-specific modeling of spatial variability across tissues and disease contexts. Through extensive benchmarking on simulated and real datasets, VISGP demonstrated superior sensitivity and robustness compared to existing approaches, while maintaining strict false discovery control. Beyond methodological innovation, VISGP revealed biologically meaningful spatial architectures—ranging from the laminar organization of the mouse olfactory bulb (*23*) to tumor–stroma interaction networks in breast cancer, squamous carcinoma, and pancreatic ductal adenocarcinoma (*26, 32*). Together, these findings establish VISGP as a generalizable statistical foundation for decoding spatial gene regulation and intercellular communication in complex tissues.

VISGP introduces two key algorithmic advances that collectively enhance both scalability and flexibility in modeling SRT data. First, by combining sparse Gaussian process approximation with variational inference (*19, 20*), VISGP circumvents the cubic computational bottleneck of conventional GP models, reducing complexity from 𝒪 (*n*^3^) to 𝒪 (*nm*^2^) while maintaining model accuracy and efficiency. The use of inducing variables allows the model to compress spatial information into a low-dimensional representation. Second, joint optimization of kernel hyperparameters through variational inference provides gene-specific adaptation of spatial covariance structures, improving sensitivity to diverse spatial patterns. Compared with existing methods such as SPARK, SpatialDE, and SPARK-X (*10–12*), which employ fixed or empirically tuned kernels, VISGP dynamically learns the optimal spatial scale (*ℓ*) and signal variance (*τ*) for each gene, yielding more accurate representation of heterogeneous tissue structure. Comparative simulation further confirmed that VISGP maintains high sensitivity and strict FDR control across distinct spatial configurations, demonstrating robust statistical performance across parameter regimes.

The performance of VISGP was first validated on the mouse olfactory bulb (MOB), a well-characterized tissue with distinct laminar organization (*23*). By accurately recovering known spatial expression layers—glomerular, mitral, and granule cell zones—VISGP demonstrated strong sensitivity to subtle spatial gradients that mirror true neuroanatomical boundaries. Compared with other approaches, VISGP produced sharper spatial delineation and higher spatial autocorrelation scores, consistent with its ability to model gene-specific covariance structures. Functional enrichment of VISGP-identified SVGs further revealed synaptic and neuronal processes, including neuron-to-neuron and asymmetric synapses, confirming that VISGP captures biologically meaningful spatial variation reflective of tissue structure rather than noise. This analysis establishes VISGP as a reliable approach for dissecting spatially organized gene programs in complex tissues. We next applied VISGP across multiple tumor contexts to evaluate its capacity to resolve spatial transcriptional heterogeneity in malignant tissues. In breast cancer, VISGP delineated two dominant spatial programs distinguishing metabolically active paracancerous regions from extracellular matrix (ECM)–remodeling tumor cores, capturing the metabolic–stromal dichotomy characteristic of invasive carcinomas (*23*). In cutaneous squamous cell carcinoma (SCC), VISGP recovered both known and novel spatial gene modules, including TNC, ITGA5, and LAMB2, which highlight an ECM– integrin axis mediating fibroblast-driven invasion and mechanical coupling (*27–29*). These findings parallel emerging evidence that the tumor stroma orchestrates invasion and immune modulation through integrin–laminin signaling (*36*). In pancreatic ductal adenocarcinoma (PDAC), VISGP identified spatially constrained ligand–receptor pairs, including TIMP1–CD63 and LAMC2–ITGB4, that define localized tumor–stroma communication niches (*32, 34, 35*). Together, these analyses reveal conserved spatial axes—ECM–receptor interaction, focal adhesion, and PI3K–Akt signaling— through which VISGP uncovers the molecular structure linking cell–matrix adhesion, metabolic remodeling, and intercellular communication across distinct tumor ecosystems.

Beyond identifying spatially variable genes, VISGP extends naturally to the inference of spatially constrained ligand–receptor interactions. There are currently three main strategies for inferring cell-to-cell communication (*42*). The first relies exclusively on single-cell RNA sequencing (scRNA-seq) to predict ligand–receptor interactions at the cell-type level, with notable tools including CellChat (*43*) and CellPhoneDB (*39*). However, these methods lack spatial context, which can lead to false positives, as most intercellular communication is restricted to spatially proximal cells. A second class of methods integrates scRNA-seq with SRT to improve spatial resolution. These approaches refine cell-type distributions using spatial data and enhance communication inference accordingly. A third class, represented by tools such as Giotto (*44*), infers communication directly from ST data. While these spatially informed approaches provide improvements, they often focus on cell-type distributions rather than the spatial expression patterns of individual genes. Notably, among the four major modes of cell-to-cell communication in the human body, three (paracrine, juxtacrine, and autocrine) require spatial proximity between interacting cells (*45*). This implies that the expression patterns of functionally interacting ligands and receptors should exhibit spatial overlap. By explicitly modeling the spatial co-expression of ligand and receptor transcripts, VISGP captures signaling relationships that are contingent on tissue structure rather than cell-type proximity. In pancreatic ductal adenocarcinoma (PDAC), this approach revealed spatially co-localized ligand–receptor pairs, including TIMP1–CD63 and LAMC2–ITGB4, which define distinct communication niches at the tumor–stroma interface (*32, 34*). Many of these interactions converge on signaling cascades such as ECM–receptor, cytokine–cytokine receptor, focal adhesion, and PI3K–Akt pathways, which collectively govern oncogenic processes including cell proliferation, adhesion, and stromal communication (*26, 40, 41*). These findings highlight how spatial modeling uncovers mechanistically grounded patterns of tumor signaling that would remain undetected in single-cell analyses lacking positional context (*39*). Importantly, several of the spatially enriched signaling pathways identified by VISGP, such as PI3K–Akt and focal adhesion, have corresponding therapeutic inhibitors in clinical evaluation, suggesting that spatially localized pathway activation could inform region-specific therapeutic vulnerabilities within the tumor microenvironment. The cross-tissue consistency of ECM–adhesion and PI3K–Akt axes underscores that spatially anchored transcriptional regulation represents a unifying principle across morphologically distinct disease contexts.

Despite its strong performance, VISGP has several limitations that suggest future directions for improvement. First, the number of inducing variables (*m*) in the sparse approximation remains a critical trade-off between computational efficiency and model fidelity. In this study, we used a fixed *m* = 20 across datasets for comparability, but adaptive strategies—such as setting *m* proportionally to the number of spatial locations or learning it via Bayesian optimization—could further enhance scalability and accuracy. Second, the current implementation employs a single Gaussian kernel to model spatial covariance. While this kernel effectively captures smooth spatial patterns, it may underrepresent complex, multi-scale, or anisotropic structures. Multi-kernel or nonstationary extensions, akin to the Cauchy combination strategy used in SPARK (*46*), may improve expressiveness but will require additional regularization to maintain tractability. Finally, integration of VISGP with complementary spatial modalities—such as spatial proteomics, imaging mass spectrometry, or multiplexed immunofluorescence—could provide richer insights into coordinated molecular and phenotypic organization. As spatial omics technologies continue to evolve toward higher resolution and multi-layer integration, VISGP offers a statistically principled foundation for scalable and interpretable spatial modeling across diverse biological systems.

## Materials and Methods

### Gaussian process model

We aim to model gene expression data of *p* genes on *n* spots measured by SRT. For a given gene, let **y** = (*y*_1_, *y*_2_, …, *y*_*n*_)^*T*^ corresponds to a gene expression vector at *n* spatial locations **X** = (**x**_1_, **x**_2_, …, **x**_*n*_)^*T*^. The coordinates of the spatial locations are typically two-dimensional, i.e. 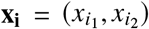. We model the expression profile of each gene individually with a Gaussian process.

A Gaussian Process (GP) represents a probability distribution over functions *f* (**x**), written as

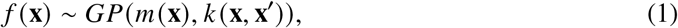

which is defined by a mean function *m* (**x**) and a covariance function *k* (**x, x**^′^). Here we assume the mean function *m* (**x**) = 0. The covariance function typically depends on some hyperparameters. In this study, we consider the exponential quadratic kernel 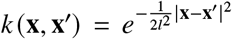 with the kernel parameter *l*.

Suppose we observe expression data 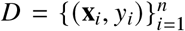 for a given gene across *n* spatial spots. Each observation *y*_*i*_ is modeled as a realization of an underlying Gaussian process with additive noise:

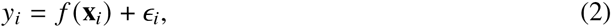

where 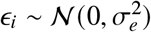. In this formulation, the input **x**_*i*_ denotes the spatial coordinate of the *i*-th spot, and the output *y*_*i*_ corresponds to the expression value of the given gene in the *i*-th spot. We rewrite the GP prior as

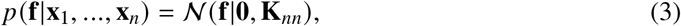

where **f** = (*f*_1_, …, *f*_*n*_)^*T*^ is a vector of function values, *f*_*i*_ = *f* (**x**_*i*_) and **K**_*nn*_ is a covariance matrix, whose entries are given by the covariance function between all pairs of spots. The joint probability model is *p* (**y, f**) = *p* (**y**|**f**) *p* (**f**), where 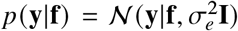 is the likelihood and *p* (**f**) is the GP prior. Then we can apply the multivariate Gaussian linear transformation rule to derive the marginal likelihood,

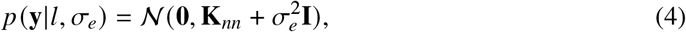

with the parameters *l, σ*_*e*_. We further propose a parameter *τ* as a scaling factor to determine the size of the covariance. Then we can rewrite the marginal likelihood of formula (4) as

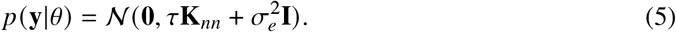

where 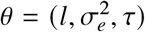. Later, in the hypothesis testing section, we show that *τ* can be interpreted as a parameter governing the variance of the spatial effect. According to Equation (5), the marginal likelihood *p* (**y** | *θ*) depends on all model parameters *l, τ*, and 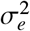. To estimate these parameters, a common approach is to maximize the log marginal likelihood log *p* (**y** | *θ*) using gradient-based optimization. Unfortunately, evaluating the objective function requires the matrix inversion 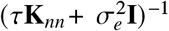 during parameter learning. For a dataset of size *n*, exact inference incurs a computational complexity of *O* (*n*^3^) and a memory cost of *O* (*n*^2^), which limits its applicability to medium or large datasets. To address this issue, a common strategy is to construct a low-rank approximation of the covariance matrix using inducing variables (*19, 47*). In this work, we adopt this approximation approach, often referred to as a sparse model (see below for details).

### Sparse Gaussian process and variational approximation

The sparse model is to build a low-rank approximation to the covariance matrix **K**_*nn*_ based on some inducing variables **u** = (*u*_1_, …, *u*_*m*_). Let the vector **u** contain values of the function *f* at some inducing points 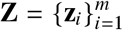 which exist in the same space as **x**, i.e. *u*_*i*_ = *f* (**z**_*i*_). The number of inducing variables *m* is required to be smaller (often significantly smaller) than the number of training data points *n*. The unknown inducing points are model parameters. We model the relationship between **u** and **f** using a multivariate Gaussian distribution, which defines our new prior. This formulation is referred to as the sparse prior, as it incorporates the inducing variables **u**, written as:

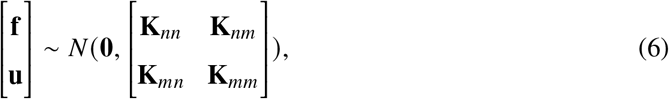

where **K**_*mm*_ is the covariance matrix evaluated between all the inducing points and 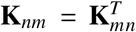 denotes the covariance matrix between all inducing points and training inputs. Upon introducing the inducing variable *u*, the marginal likelihood can no longer be expressed in the form of Eq. (5). Consequently, we turn our attention to the posterior distribution *p* (**f, u**|**y**),

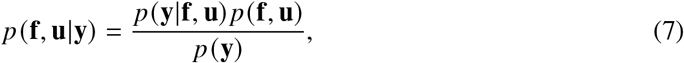

which is a joint distribution over the random variable vector **f** and **u**. Note that this formula also mentions the marginal likelihood *p* (**y**), which contains the *n* × *n* covariance matrix **K**_*nn*_. Therefore, it remains inconvenient for us to carry out a precise derivation.

To solve the above problem, we employ a new distribution *q*(**f, u**), referred to as the variational distribution, to approximate the true posterior *p* (**f, u** | **y**). Instead of finding an exact GP model, we minimize the divergence between the exact posterior GP and the variational distribution. The inducing points **Z** become variational parameters, which are learned together with the model parameters to minimize this distance. Let us suppose the variational distribution *q*(**f, u**) is a joint distribution with the following factorization:

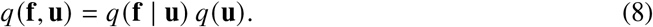

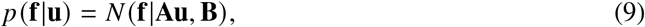

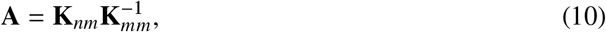

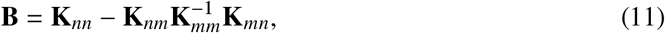

where *p* (**f** |**u**) is obtained by applying the conditional rule of multivariate Gaussian distributions to the sparse prior *p* (**f, u**), and *q*(**u**) is a multivariate Gaussian distribution, i.e.

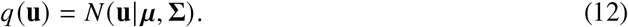

Since log *p* (**y**) is difficult to represent directly using the inducing variable *u*, it can be approximated by the variational lower bound (ELBO). By maximizing the ELBO to learn the model parameters, we can ultimately obtain our model in Eq. (5). The ELBO is derived as follows:

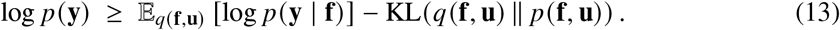

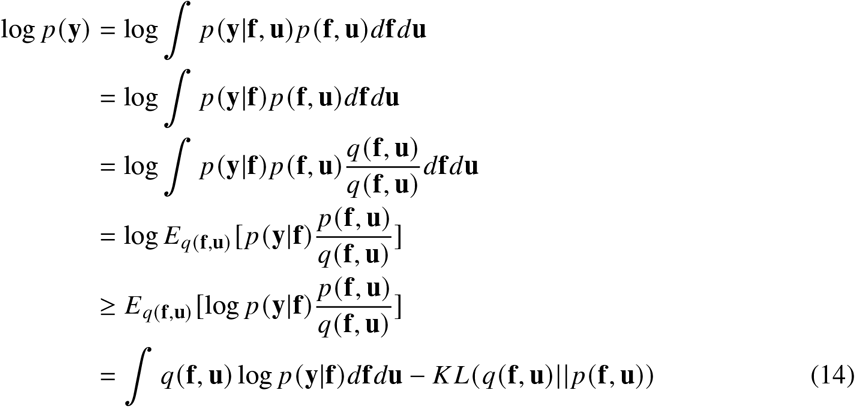

The resulting ELBO consists of two parts: a likelihood term and a KL divergence term. We now manipulate the likelihood term further:

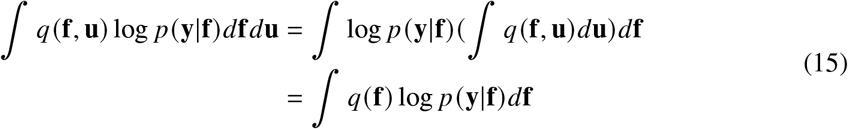

It is important to note that *q*(**f**) represents the distribution of the random variable vector **f**, and it is distinct from the marginal variational distribution *q*(**u**) = *N* (**u**| ***μ*, Σ**). Since the joint variational distribution *q*(**f, u**) is not defined as a multivariate Gaussian distribution, we cannot directly obtain the marginal distribution *q*(**f**) from it. So, it is necessary to explicitly derive the analytical expression for *q*(**f**).

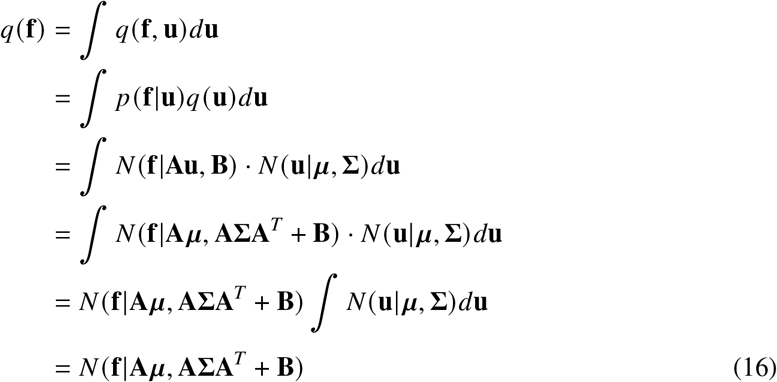

Now we derive the analytical expression for the KL term in the *ELBO*:

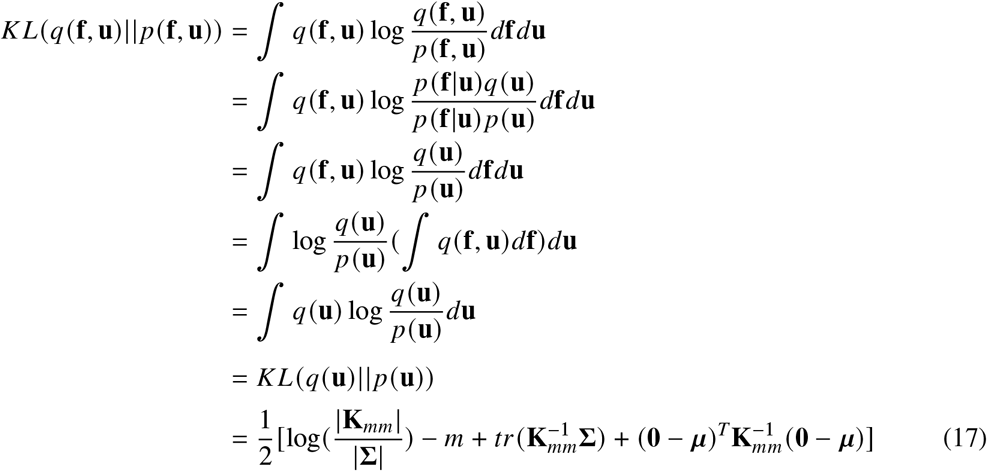

Finally, we can get the objective function:

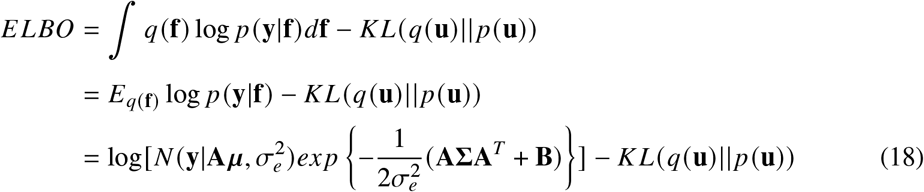

The analytical expression for the *ELBO* depends on all our model parameters as its arguments. To determine the optimal values for these parameters, we maximize the *ELBO* using gradient descent.

### Hypothesis testing

We assess spatially variable genes (SVGs) through a hypothesis testing framework, where the null hypothesis posits the absence of spatial structure in gene expression. From the preceding derivation, the distribution of gene expression values **y** across spatial locations is given by Eq. (5):

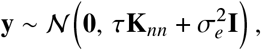

with *τ* denoting the variance component attributable to spatial dependence. Thus, the presence of a spatial expression pattern can be formally evaluated by testing the null hypothesis

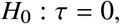

which corresponds to the case where no spatial variation is explained by the covariance structure. We compute a p-value using the Davies method (*48*), based on the score statistic

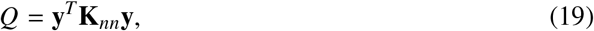

which follows a mixture of *χ*^2^ distributions (see Supplementary Materials for derivation details). Genes are then selected as spatially variable if their q-value (adjusted p-value, FDR (*49*)) is less than 0.05, and are consequently considered SVGs.

### Datasets and preprocessing

We obtained mouse olfactory bulb and human breast cancer data from the SRT Research database (https://www.spatialresearch.org/). These datasets consist of gene expression measurements derived from read counts across discrete spatial locations, referred to as spots. Our analysis included the MOB replicate 9 dataset, comprising measurements of 15,284 genes across 237 spots, and the BC layer 2 dataset, comprising 14,789 genes across 251 spots. Genes with fewer than ten total counts were filtered out, and only spots with a minimum of ten total read counts were retained for subsequent analysis. We analyzed 11,428 genes across 237 spots in the mouse olfactory bulb dataset and 9,907 genes across 251 spots in the human breast cancer dataset. Human squamous cell carcinoma and pancreatic ductal adenocarcinoma data were obtained from the Gene Expression Omnibus (https://www.ncbi.nlm.nih.gov/geo). For human squamous cell carcinoma, we selected Patient 5, replicate 2, which initially comprised 17,690 genes measured in 1,934 spots. Based on criteria described in the original study, a subset of 521 spots was retained for downstream analysis. 25754 genes across 428 spots were analysised in pancreatic ductal adenocarcinoma. Prior to statistical analysis, all count data were normalized using sklearn.preprocessing.scale to achieve zero mean and unit variance.

### Comparison of methods

We compared VISGP with three established methods for identifying genes with spatial expression patterns: SpatialDE (v.1.1.3), SPARK (v.1.1.1), and SPARK-X (v.1.1.1). SpatialDE employs a multivariate normal model to assess the spatial variability of gene expression, using likelihood ratio tests for significance evaluation. In this framework, a Gaussian kernel is selected via the Bayesian information criterion, and its hyperparameter is optimized from a predefined set of values. Building on SpatialDE, SPARK adopts a spatial generalized linear mixed model that incorporates multiple spatial kernels, enabling direct modeling of count-based spatial transcriptomic data. SPARK-X further extends the framework by adopting a nonparametric covariance-testing approach that supports a variety of spatial kernels, thereby enhancing scalability and performance on large-scale, sparse spatial transcriptomics datasets. For SpatialDE, SPARK, and SPARK-X, we applied consistent pre-processing criteria to filter genes and spots, ensuring that all methods performed SVG analysis on identical datasets. Following the guidelines and example workflows provided in their respective GitHub repositories, we evaluated each method on a simulated dataset, as well as on three experimental datasets: mouse olfactory bulb, human breast cancer, and human squamous cell carcinoma.

### Statistical Analysis

#### TPR and TNR

The true positive rate 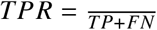 is used to measure the method’s sensitivity. A method with a higher sensitivity has a lower type II error rate. The true negative rate 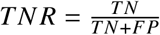 could measure the specificity of the method. A method with a higher specificity has a lower type I error rate.

#### Moran’s I Statistic

Moran’s I statistic is a widely used index for assessing the degree of spatial autocorrelation. The Moran’s I statistic value is distributed in [−1, 1]. When the value is closer to 1, it means that the attribute values on the spatial points are more aggregated into a specific pattern; when the value is more comparable to −1, it means that the attribute values on the spatial points are more scattered; and when the value is equal to 0, it means that the attribute values of the spatial points are completely randomly distributed. We used the R package spdep (v.1.4.1) to compute the Moran’s I statistic.

#### Gene Set Enrichment Analysis

We performed GO term enrichment analysis using the R package clusterProfiler (v.4.14.6). In the package, we used the default “BH” method for multiple testing corrections and screened GO terms with FDR less than 0.05 as significantly enriched. The results showed the top 10 most enriched GO terms for the three categories of GO terms (cellular component, molecular function, and biological process), respectively.

## Data Availability

All data produced are available online at

## Acknowledgments

We would like to thank Prof. Xiang Zhou and his team for their valuable discussions on the statistical model, which greatly improved the quality of this work.

## Author contribution

Conceptualization: JLH, JL, YRW, ZCW

Methodology: JLH, YRW

Investigation: ZCW, LQX, YRW

Visualization: ZCW, LQX

Supervision: JLH

Writing—original draft: ZCW, LQX, YRW

Writing—review & editing: JLH, JL, TW, YTW, XQS

## Data and materials availability

All data analyzed in this paper can be obtained from the original study. Specifically, the mouse olfactory bulb dataset and human breast cancer dataset are collected from the SRT Research website (https://www.spatialresearch.org). The human squamous cell carcinoma dataset and pancreatic ductal adenocarcinoma are obtained from the Gene Expression Omnibus under accession number GSE144240 and GSE111672 respectively. The VISGP algorithm is implemented in the toolkit package “scbean” (≥ v0.5.0) in the PyPI repository, and the source code can also be freely accessible at https://github.com/jhu99/scbean.

## Supplementary Materials

Supplementary Text

Figs. S1 to S9

## Supplementary Materials for

## Supplementary Text

### Derivation details of score statistic obeying mixture *χ*^2^ distribution

Under the null hypothesis *τ* = 0 and thus,

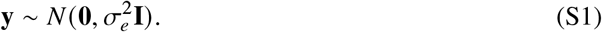

If we premultiply **y** by a matrix 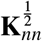, we can get

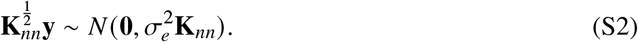

We decompose the covariance matrix 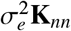:

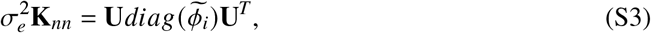

where 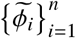 are the eigenvalues of matrix 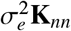, and **U** is a matrix composed of corresponding eigenvectors. According to the properties of matrix decomposition, we can know that **UU**^*T*^ = **I**. Then, based on formula (S2), we premultiply matrix **U**^*T*^ on the left,

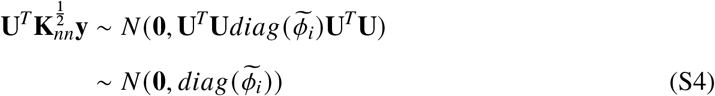

Finally, we get our score statistic *Q* and its distribution,

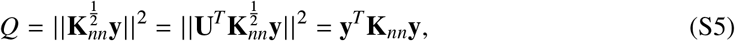

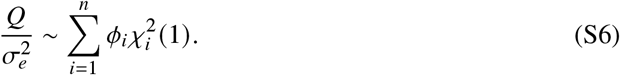

where {*ϕ*_1_, …, *ϕ*_*n*_} are the eigenvalues of **K**_*nn*_.

The above derivation is exact whenever 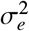 is known. However, in practice, 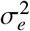 is not known and needs to be estimated from the data. Here, 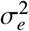 is replaced with its maximum likelihood estimate under the null hypothesis,

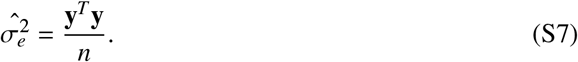

The statistic *Q* and 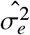 are in fact dependent random variables. Therefore, the assumed distribution of 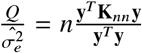 does not hold when substituting 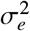 with its estimate, 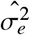. Through the derivation of formula (S6), we can effectively calculate the p-value for the distribution of the statistic 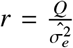 calculated from the observed data.

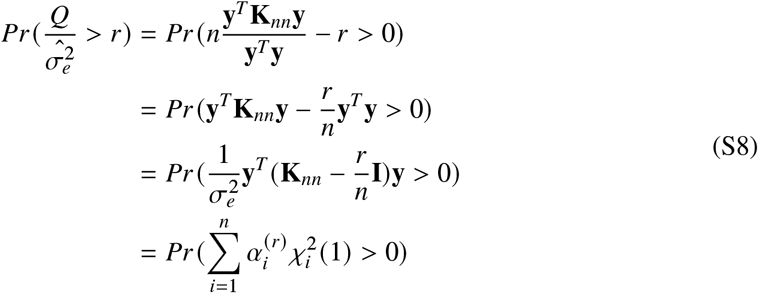

Similar to the previous derivation, 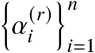 represent the eigenvalues of matrix 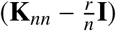.

## Supplementary Figures

**Figure S1:**
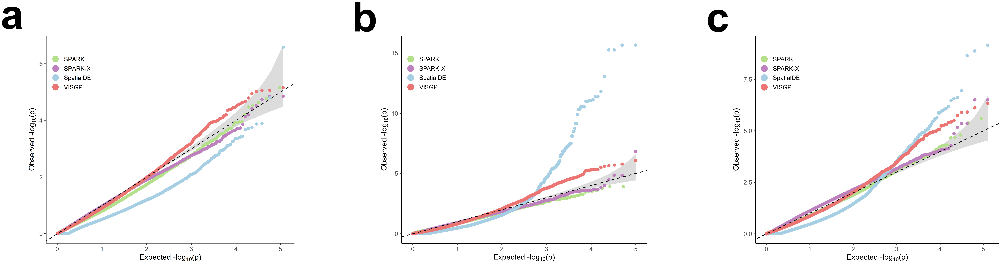
Null quantile–quantile plots across datasets. Quantile–quantile plots of the observed − log_10_(*p*) from different spatial methods are plotted against the expected − log_10_(*p*) under the null in the permuted data. p-values were combined across ten permutation replicates. Compared methods include SPARK (green), SPARK-X (purple), SpatialDE (blue), and VISGP (red). (a), (b), and (c) correspond to the mouse olfactory bulb, the human breast cancer, and the human squamous cell carcinoma datasets, respectively.

**Figure S2:**
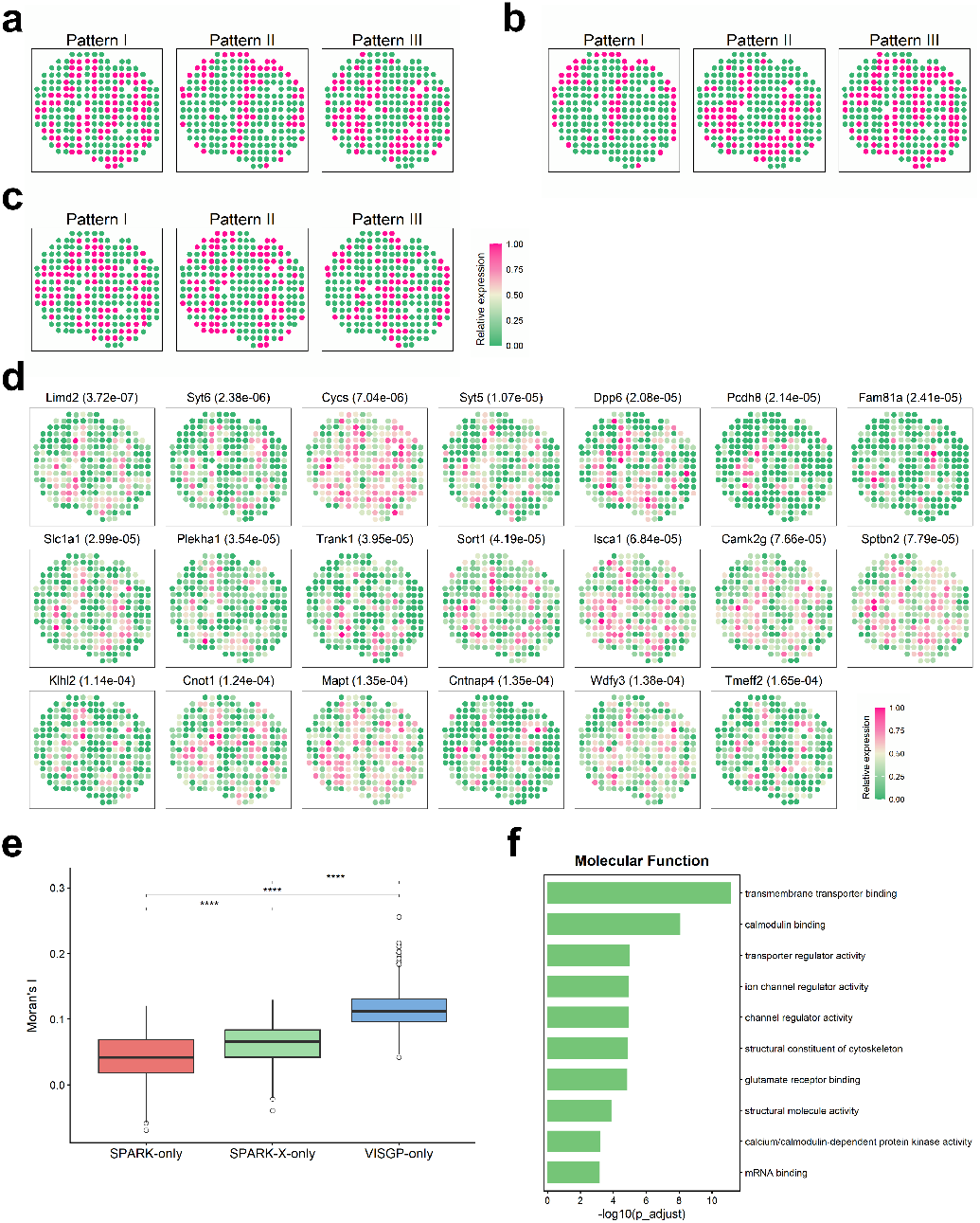
Supplementary of the mouse olfactory bulb dataset. (a), (b), and (c) show three spatial patterns by K-means clustering according to the SVGs identified only by VISGP, SPARK and SPARK-X, respectively. (d) Spatial expression pattern for some SVGs identified only by VISGP. (e) Boxplot of Moran’s I statistics of SVGs identified by SPARK only, SPARK-X only and VISGP only. (f) Bar plot for GO molecular function enrichment analysis on SVGs obtained by VISGP based on an FDR cutoff of 0.05.

**Figure S3:**
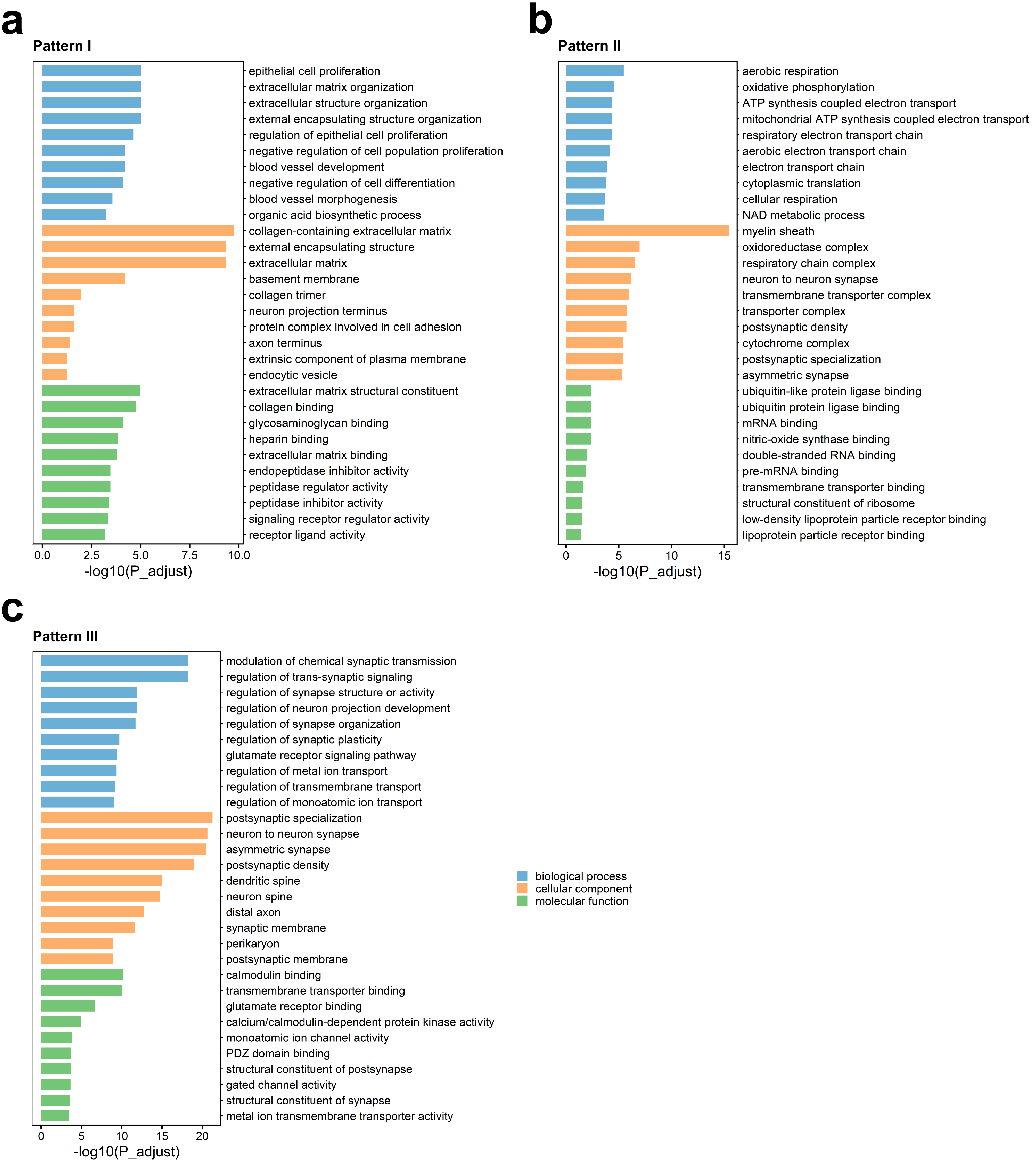
Go enrichment analysis of SVGs identified by VISGP in the mouse olfactory bulb dataset. (a–c) Bar plots showing GO enrichment analysis of SVGs from three spatial patterns: (a) Pattern I, (b) Pattern II, and (c) Pattern III.

**Figure S4:**
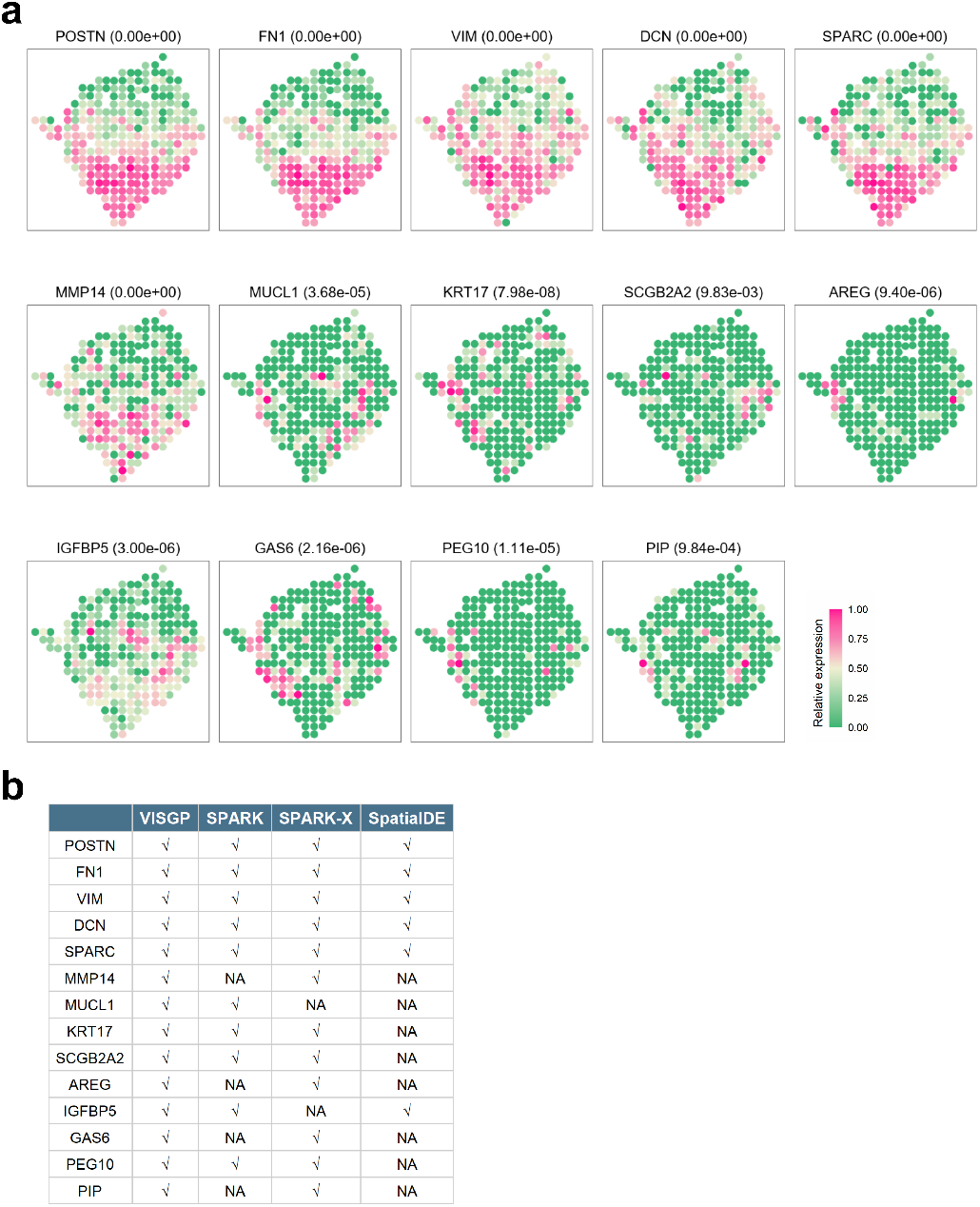
SVGs identified by VISGP and comparison with other methods. (a) Spatial expression pattern for these 14 genes. VISGP recognizes all of them as SVGs along with their FDR (in parentheses). (b) Comparison of detection results across four methods (VISGP, SPARK, SPARK-X, and SpatialDE); a checkmark indicates successful detection of the gene related to cancer progression.

**Figure S5:**
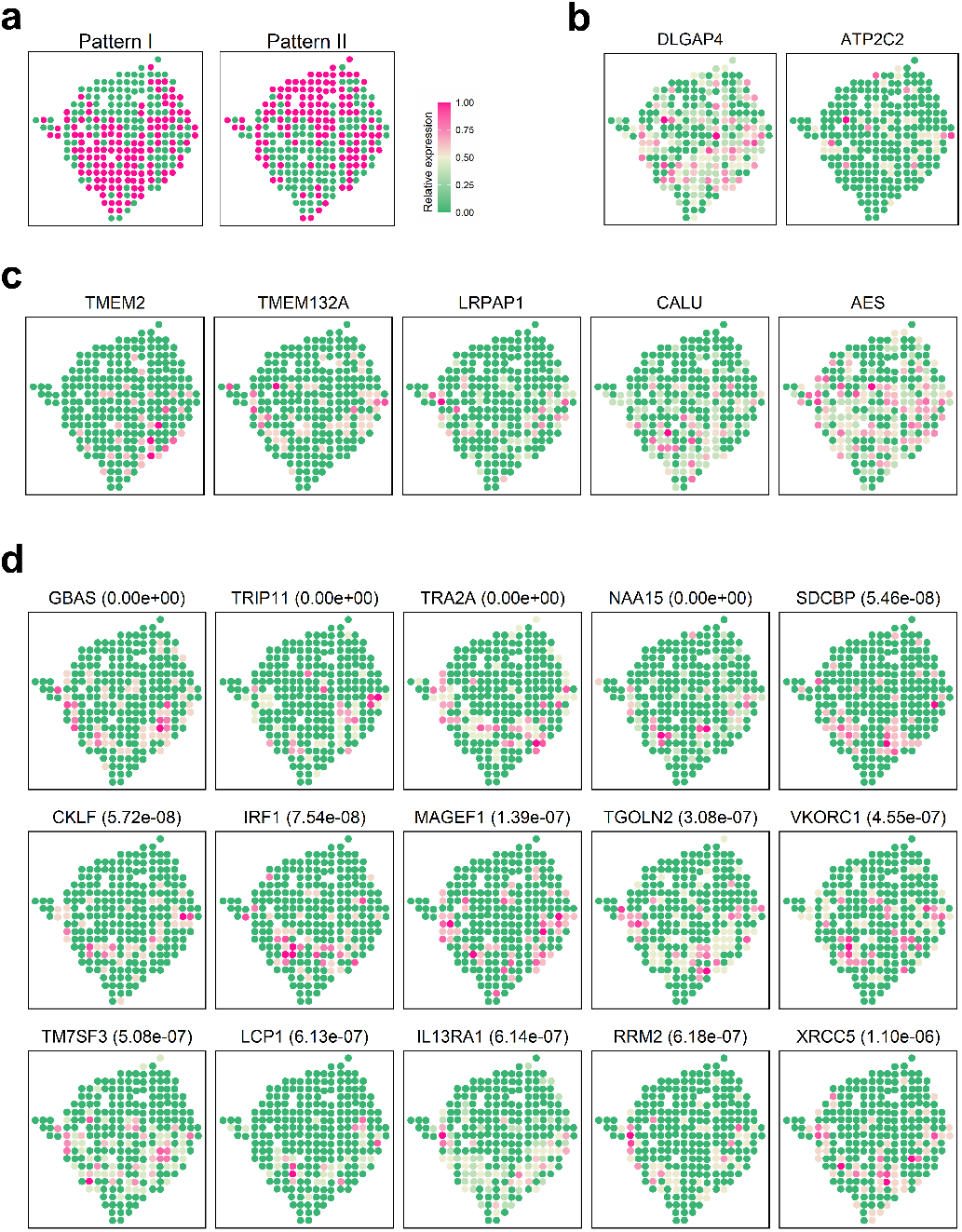
Supplementary of the human breast cancer dataset. (a) According to the SVGs identified only by VISGP, two main spatial expression patterns are summarized. (b) and (c) show spatial expression pattern for SVGs identified by SPARK only and SPARK-X only. (d) Spatial expression pattern for some SVGs identified only by VISGP.

**Figure S6:**
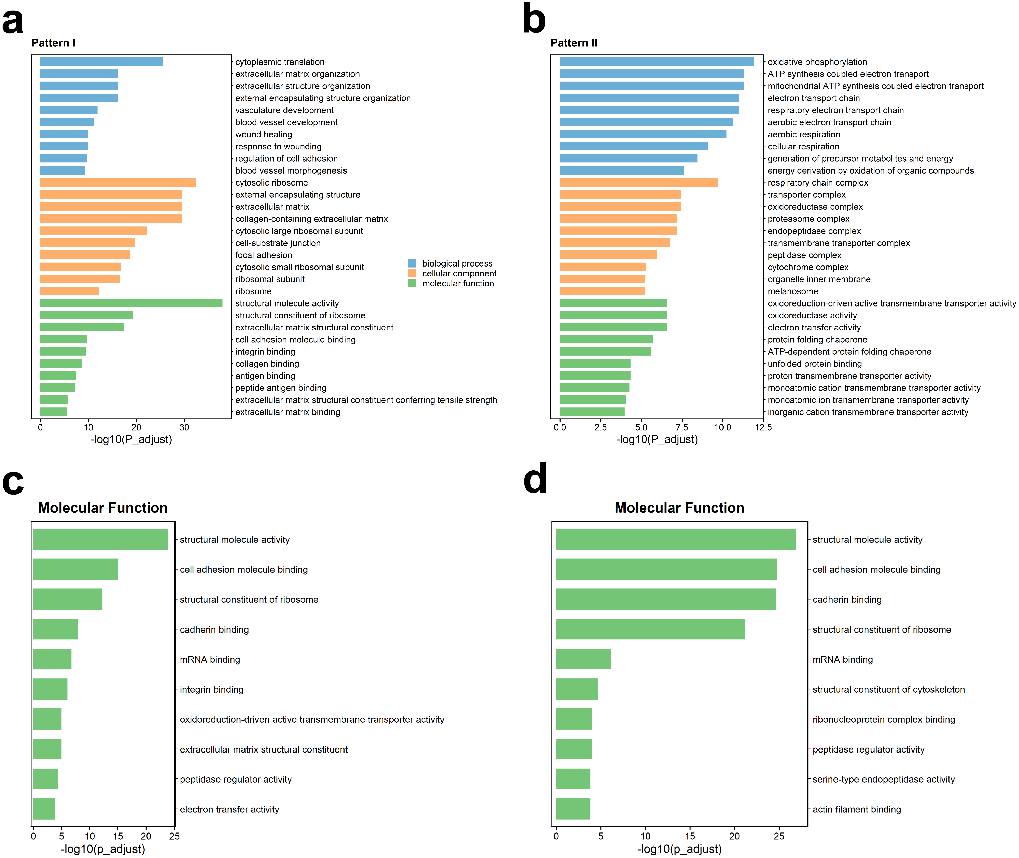
Go enrichment analysis of SVGs identified by VISGP in the human breast cancer dataset and the human squamous cell carcinoma dataset. (a) and (b) Bar plot for GO enrichment analysis on SVGs belonging to Pattern I or Pattern II in the human breast cancer dataset. (c–d) Bar plots showing GO molecular function enrichment of SVGs obtained by VISGP, with panel (c) corresponding to the human breast cancer dataset and panel (d) corresponding to the human squamous cell carcinoma dataset.

**Figure S7:**
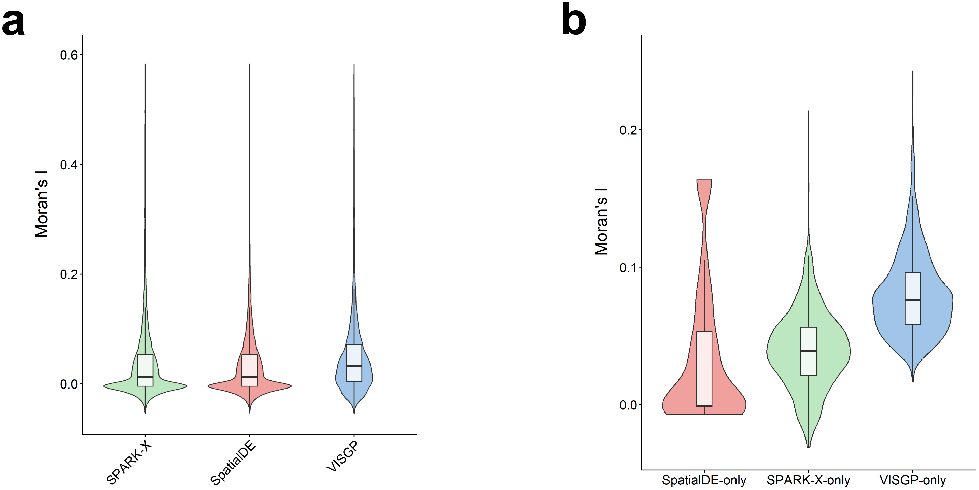
Moran’s I spatial autocorrelation statistics test in human squamous cell carcinoma. (a) Violin-plot of Moran’s I statistics of SVGs identified by SPARK-X, SpatialDE and VISGP. (b) Violin-plot of Moran’s I statistics of SVGs identified by SpatialDE only, SPARK-X only and VISGP only.

**Figure S8:**
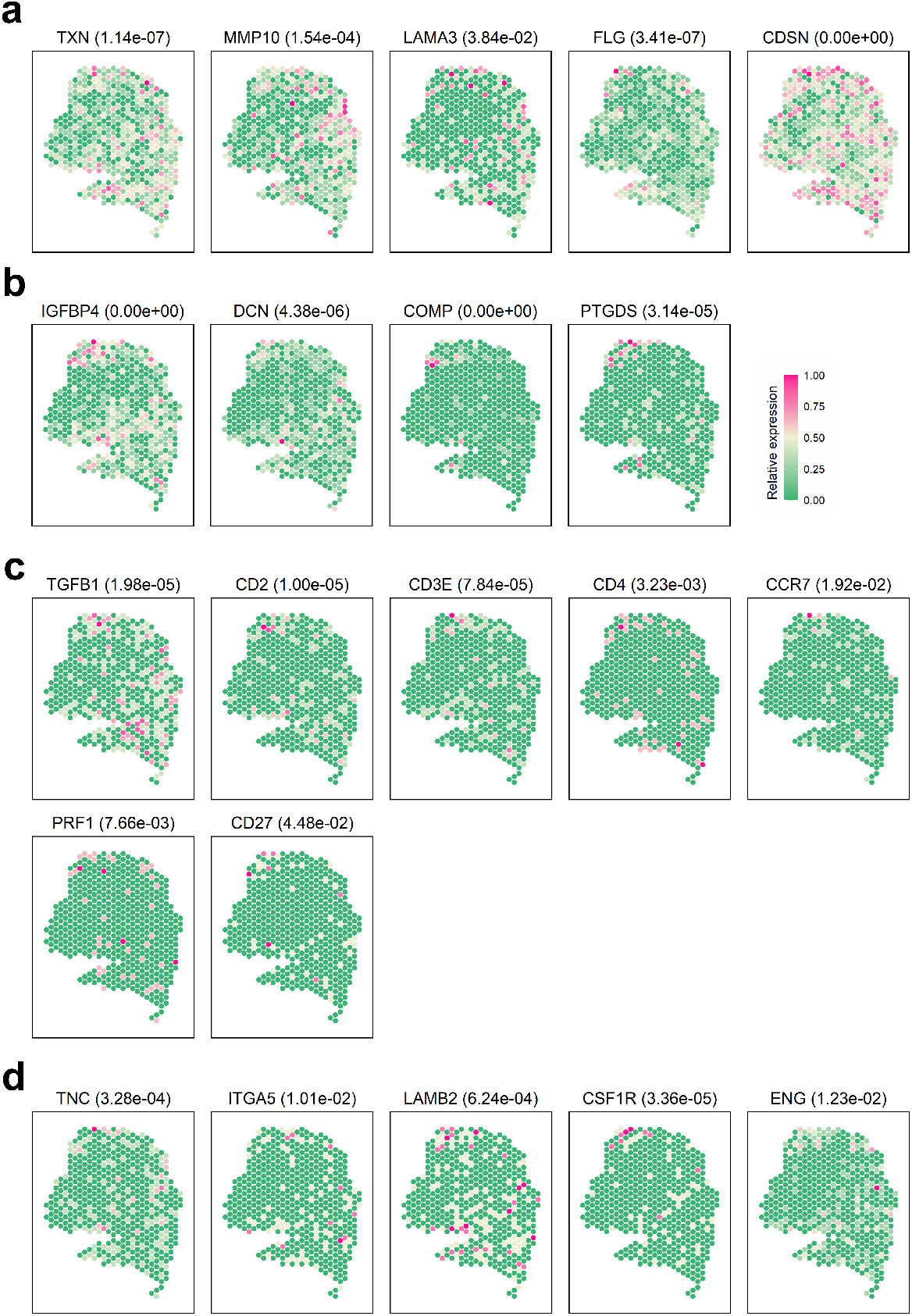
Spatial expression pattern for some SVGs identified by VISGP in the human squamous cell carcinoma dataset. (a–b) SVGs display distinct spatial localizations, with (a) showing tumor-associated SVGs and (b) showing SVGs enriched in the non-tumor adjacent stroma. (c) Some SVGs include immunosuppression-associated genes (TGFB1), T cell marker genes (CD2, CD3E, CD4, CCR7), and genes related to cytotoxic (PRF1) or co-stimulatory (CD27) functions. (d) Some SVGs are identified only by VISGP.

**Figure S9:**
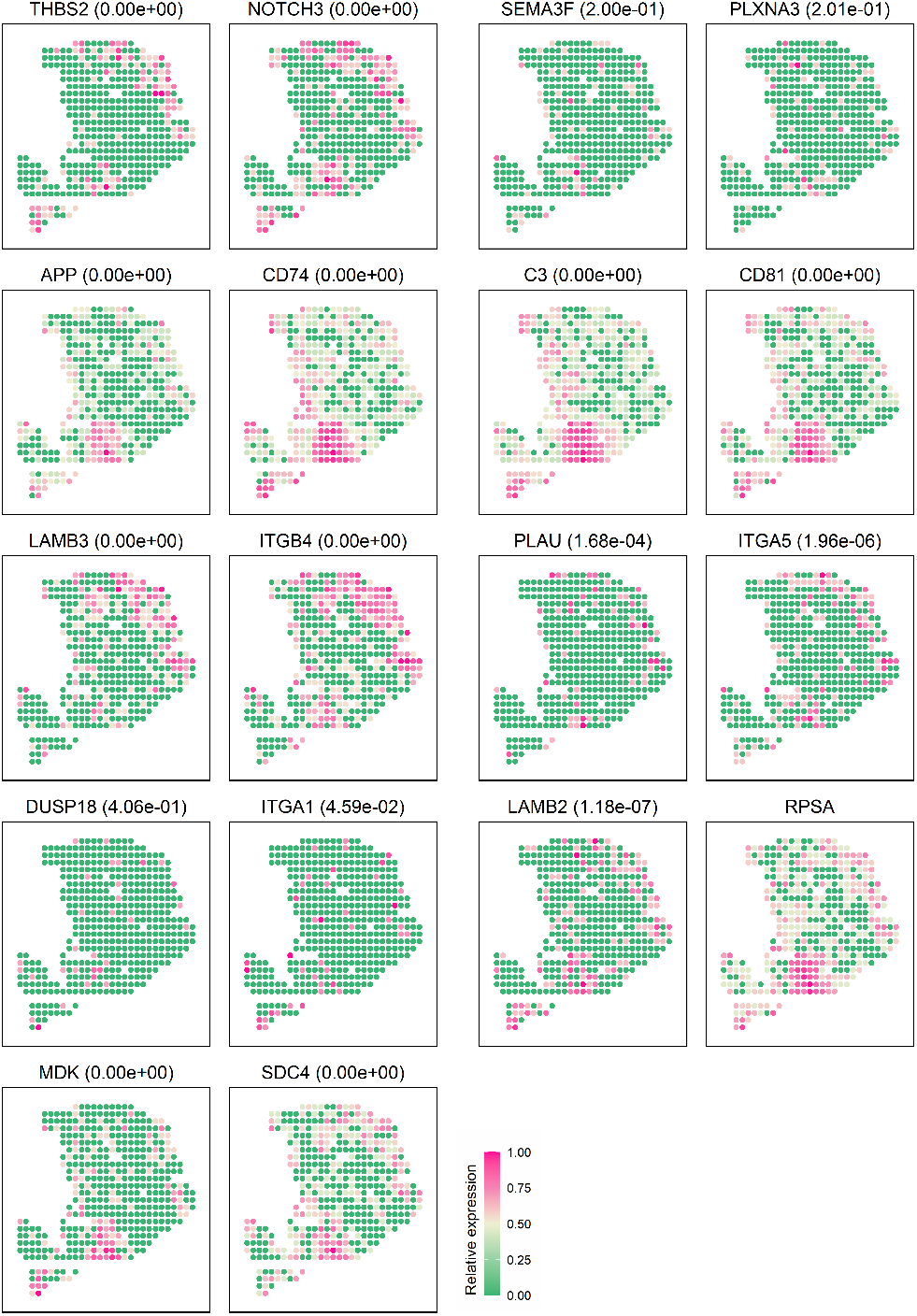
Heatmaps of ligand-receptor pairs detected by VISGP that exhibit similar spatial expression distributions.

